# Increasing Efficiency, Persistent Burden: Longitudinal Analysis of EHR Use and After-Hours Work in Emergency Medicine Residency

**DOI:** 10.64898/2026.05.19.26353524

**Authors:** Carl Preiksaitis, Josh Hughes, Mark Iscoe, Michael Makutonin, Ashley Rider, Edward R. Melnick, Christian Rose

**Affiliations:** Department of Emergency Medicine, Stanford University; Department of Emergency Medicine, Yale University.

## Abstract

**Objectives:** Electronic Health Records (EHRs) impose a significant time burden on physicians, often requiring work to be completed outside of scheduled hours. While this burden is well-documented, how it evolves throughout emergency medicine (EM) residency remains poorly understood. This study aimed to quantify EHR usage patterns, analyze the composition of after-shift work, and characterize the development of EHR efficiency across EM training.

**Methods:** We conducted a retrospective cohort study of EM residents (postgraduate year [PGY] 1–4) using 5.5 years of EHR audit log data (2020–2025) at a single academic institution. We analyzed EHR time per new patient encounter, stratified by postgraduate year, and categorized activities into domains such as documentation, chart review, and orders. EHR work was measured both during and after scheduled shifts.

**Results:** The analysis included 144 unique residents and 167,010 new patient encounters across 15,386 shifts. Encounter-attributed EHR time per encounter decreased by 52% from PGY-1 to PGY-4 (median 19.9 to 9.6 minutes, p<0.001), despite an 86% increase in patient volume per shift (median 7 to 13 encounters). This efficiency gain was driven primarily by a 69% reduction in documentation time (9.3 to 2.9 minutes), accompanied by shorter notes. After-shift work (EHR activity after the 9-hour clinical shift) was present in 89.9–94.4% of encounters. At the shift level, combined after-shift EHR time (encounter-attributed plus tracking board) was a median of 64.2 minutes per shift for PGY-1 and 104.2 minutes for PGY-4. Shift-level tracking board activity dominated the after-shift burden and increased with training (median 40.2 to 79.0 minutes per shift from PGY-1 to PGY-4).

**Conclusions:** EM residents achieve substantial gains in on-shift EHR efficiency, with the largest reductions observed in documentation time, accompanied by shorter notes and faster input speed. However, a persistent after-hours workload, dominated by administrative and patient flow tasks, suggests that (at least at this single institution) system-level factors—not just individual skill—may contribute to this pattern. Monitoring these objective EHR metrics may help programs identify struggling learners and evaluate the impact of interventions aimed at improving resident well-being and workflow efficiency.

## INTRODUCTION

Electronic health records (EHRs) have improved safety surveillance, information access, and care coordination, but they also impose substantial time costs on clinicians.^1-3^ In the emergency department (ED), direct observation studies show that physicians divide their time roughly equally between computer-related activities and direct patient interaction, and emerging audit-log benchmarks demonstrate that per-encounter EHR time varies substantially with patient acuity and clinical factors.^4,5^ In ambulatory practice, for every direct hour of patient care, physicians spend nearly two additional hours on EHR and desk work, with additional work commonly occurring outside of scheduled hours.^6-8^ Concurrently, standardized audit-log metrics have matured, enabling reproducible, task-level measurement of EHR use across settings.^9,10^

The ED is a particularly demanding environment for digital work: high acuity, frequent interruptions, and rapid decision cycles intensify tensions between documentation and bedside care.^11^ Time-motion studies show that EHR implementations and order entry tools shift clinicians’ task allocation toward computer use, and post-implementation efficiency losses may persist without targeted workflow redesign.^12,13^

For trainees, these demands intersect with learning curves. Prior work in emergency medicine (EM) estimated that residents spend approximately 2,171 hours—about 7.7 work months—interacting with the EHR over a 3-year program, with chart review and documentation comprising the largest shares.^14^ Beyond total volume, after-hours EHR work (often termed “pajama time”) has emerged as a salient burden associated with well-being concerns; in a large longitudinal resident cohort, per-patient EHR time decreased over 12 months of the academic year while the proportion of after-hours time did not, suggesting structural contributors to when work occurs.^15^

However, postgraduate year (PGY)-specific, encounter-level characterization of *what* residents do in the EHR within a shift-based EM context and how the composition of activities (e.g., documentation, review, orders) changes with seniority remains limited. In addition, the mechanisms that drive after-shift work in EM training are incompletely described. To address these gaps, we used comprehensive audit-log data from a four-year EM residency to (1) quantify total EHR time per encounter and the distribution of within-shift versus after-shift work across PGY levels; (2) decompose resident EHR activity by task category; and (3) examine documentation-focused efficiency metrics to clarify mechanisms of change with training. We hypothesized that on-shift efficiency would improve with seniority, whereas a residual after-shift burden would persist.

## METHODS

### Study design and setting

We performed a retrospective cohort study using EHR audit log data for adult (Age ≥ 18) patients from a single clinical site for a large, four-year academic EM residency. The observation window extended from January 1, 2020 through July 31, 2025. The institutional review board approved the study (Protocol # 69107).

### Study population

The source population included all EM residents PGY1–4 who worked clinical shifts in the adult ED during the study period. This design allowed both cross-sectional comparisons across training levels and within-individual assessment for residents with longitudinal observation.

### Encounter attribution and cohort construction

The analytic unit was a resident-encounter-shift record. Encounter inclusion and resident assignment were derived directly from the EHR’s Treatment Team table to align with the institution’s standardized method for attributing clinical volume. Under this institutional logic, an encounter is strictly attributed to the first resident who formally signs up for the patient. At our institution, documentation is shared across the team; however, the first resident to sign up typically acts as the primary provider and composes the comprehensive note (history of present illness, review of systems, physical examination, and medical decision making). Because the institutional logic attributes the encounter only to this primary provider, residents taking over a patient during transitions of care (sign-outs) do not receive encounter credit in this dataset. This naturally focuses the cohort on primary patient encounters where the resident is responsible for the bulk of the documentation.

To further improve comparability and reduce role-related confounding, the primary analysis applied two additional exclusion criteria. First, we excluded purely administrative and supervisory shifts where senior residents oversee multiple teams and do little primary patient care. Second, we excluded senior resident records in encounters where a senior resident (PGY3–4) and a junior resident (PGY1–2) were simultaneously co-credited on the Treatment Team for the same patient. In these instances, the senior’s role is primarily supervisory, while the junior resident acts as the principal documenter.

### Data sources and variable derivation

Three Epic EHR data sources (Epic Systems Corporation, Verona, WI) were used in combination: event-level audit logs containing timestamped user interactions; aggregated active-use logs that quantify the duration of active EHR interaction; and a documentation attribution table that provides note-level metadata including total character counts and entry method.^16^ Resident schedules from the institutional scheduling system were used to define exact shift boundaries.

Raw audit-log events were harmonized and aggregated into clinically meaningful categories adapted from prior work.^4^ The taxonomy comprises *Notes* (documentation), *Tracking* (ED tracking board and patient-flow activities), *Review* (chart, laboratory, and imaging review), *Orders* (computerized provider order entry), *Disposition* (admission, discharge, outpatient prescriptions and referrals, and the generation of discharge instructions), *Inbox* (messaging and in-basket actions), and *Other* (miscellaneous or uncategorized events). Active EHR time was defined as the cumulative duration (in seconds) of user interaction derived from Epic’s active-use logs using the vendor’s standard inactivity threshold (5-second timeout). For each resident-encounter-shift, we calculated total active EHR time and time within each activity category.

From the documentation attribution table, we extracted note length (total characters) and the composition of characters by entry method: manual keyboard entry, dot phrase tools (SmartPhrases), voice recognition (dictation), and system-generated content (for example, SmartText, SmartLinks, or imported/copied elements). As an efficiency proxy, documentation speed was calculated as manually entered or dictated characters per minute of active documentation time.

Scribe utilization was identified at the encounter level based on audit log activity. An encounter was classified as scribed if a user with the Epic role assigned to ED scribes had ≥ 2 minutes of note-writing activity on the same encounter. This duration threshold was selected based on empiric distributions of scribe activity to distinguish meaningful documentation assistance from incidental chart access or brief addenda.

### Shift definitions and activity normalization

Residents are scheduled for nine-hour clinical shifts. The first eight hours are dedicated to direct patient care; the final hour is allocated for care transitions, sign-out, documentation, and other end-of-shift wrap-up tasks. Because our objective was to characterize after-shift EHR work as activity occurring outside any scheduled clinical time, we defined the on-shift period as the full nine-hour scheduled shift and after-shift activity as work occurring after the scheduled shift end through 48 hours post-shift, an interval chosen to capture the majority of instances of delayed documentation completion.

Tracking board time—time spent viewing the ED patient tracking board, census screen, and patient flow management displays—was analyzed separately as a shift-level metric rather than distributed across encounters because 97% of tracking activity in the audit logs was not attributed to a specific patient encounter, consistent with the nature of tracking board use as a general situational awareness and patient flow management tool. Tracking board activity in our environment was dominated (>99%) by interactions with the ED Track Board (Epic Activity ID 34798), which functions as a real-time departmental dashboard. Residents use it to monitor resulted studies, bed assignments, patient flow, and disposition status across their patient panel. In our ED, this dashboard typically remains open as a persistent secondary workspace during clinical work. Brief documentation updates are often input via the ED Track Board interface, however, this *Notes* activity is indistinguishable from other *Tracking* activities in the audit log. Encounter-attributed EHR measures were normalized per patient encounter, while tracking board time was reported per shift.

### Outcomes

Main outcomes were total active EHR time per encounter (excluding tracking board time, which was analyzed separately as a shift-level metric) and the distribution of on-shift versus after-shift time across PGY levels. Secondary outcomes included the composition of EHR activity by category and documentation features (note length, entry-method composition, and documentation speed), shift-level tracking board time, and per-shift totals of after-shift EHR work.

### Statistical analysis

We described resident characteristics and EHR workload using medians with interquartile ranges for continuous variables and counts with percentages for categorical variables. Because encounters are nested within residents, we aggregated EHR-time measures to the resident level (one median per resident per PGY year) before formal hypothesis testing. This cluster-summary approach treats each resident as a single observation within each training year, eliminating the inflated precision that would result from treating encounters as independent. Because time-based measures are typically right-skewed, we used Kruskal-Wallis tests on resident-level medians to compare PGY levels and, when overall tests were significant, Mann-Whitney U tests for pairwise comparisons. Two-sided p values < 0.05 were considered statistically significant. Analyses were conducted in Python 3.11 using pandas and scipy.

## RESULTS

### Cohort characteristics

The study included 144 unique EM residents, of whom 115 (79.9%) contributed data in more than one training year and 52 (36.1%) had data across all four PGY levels. After applying primary inclusion and exclusion criteria, the analytic dataset contained 167,010 resident-encounter pairs across 15,386 shifts: 13,521 (8.1%) PGY-1, 45,072 (27.0%) PGY-2, 68,998 (41.3%) PGY-3, and 39,419 (23.6%) PGY-4 encounters.

### Resident and patient demographics

Residents were 54.7% male and 45.3% female, with a mean age of 29.2 years at the start of training (SD 3.2, range 25–45). Racial distribution was 69.6% White, 17.6% Asian, 8.8% Other, and 4.1% Black or African American; 8.8% identified as Hispanic/Latino.

Table 1 displays the clinical characteristics of encounters stratified by PGY year. The median ESI triage level was 3 (IQR 2–3) across all training levels. PGY-1 residents cared for patients with the lowest acuity mix (28.7% ESI 1–2) and the lowest admission rate (32.7%), while PGY-2–4 residents saw a comparable acuity mix (30.2–33.4% ESI 1–2) with admission rates of 35.3–35.8%. Median ED length of stay was similar across PGY levels (366–372 minutes), and median encounters per shift increased from 7 in PGY-1 to 13 in PGY-4 (p<0.001).

**Table 1:** Clinical Characteristics by Training Year.

| **Metric** | **PGY-1** | **PGY-2** | **PGY-3** | **PGY-4** | **p-value** |
| --- | --- | --- | --- | --- | --- |
| N encounters | 13,521 | 45,072 | 68,998 | 39,419 | — |
| Median ESI (IQR) | 3 (2–3) | 3 (2–3) | 3 (2–3) | 3 (2–3) | <0.001 |
| % ESI 1–2 (high acuity) | 28.7% | 33.3% | 33.4% | 30.2% | <0.001 |
| % Admitted | 32.7% | 35.3% | 35.4% | 35.8% | <0.001 |
| % Discharged | 61.0% | 58.0% | 58.0% | 57.4% | <0.001 |
| Median ED LOS, min (IQR) | 366 (257–530) | 372 (255–544) | 372 (259–540) | 367 (256–532) | <0.001 |

### Overall EHR time burden and activity composition

Encounter-attributed EHR time patterns varied across training levels (Figure 1). Median total active EHR time per encounter (excluding tracking board time, reported separately at the shift level) decreased from 19.9 minutes in PGY-1 to 9.6 minutes in PGY-4, a 52% reduction (p<0.001). Documentation time represented the largest component of encounter-attributed EHR work at all training levels. Median documentation time per encounter decreased from 9.3 minutes in PGY-1 to 2.9 minutes in PGY-4 (69% reduction). Order time declined from 2.9 to 1.7 minutes (41% reduction), and chart review time decreased from 3.6 to 2.7 minutes (25% reduction). Group-level differences across PGY were statistically significant for total time and for each activity category (p<0.001, Kruskal-Wallis on resident-level medians); attenuation of selected pairwise comparisons after clustering is reported in the pairwise comparisons below.

**Figure 1:**
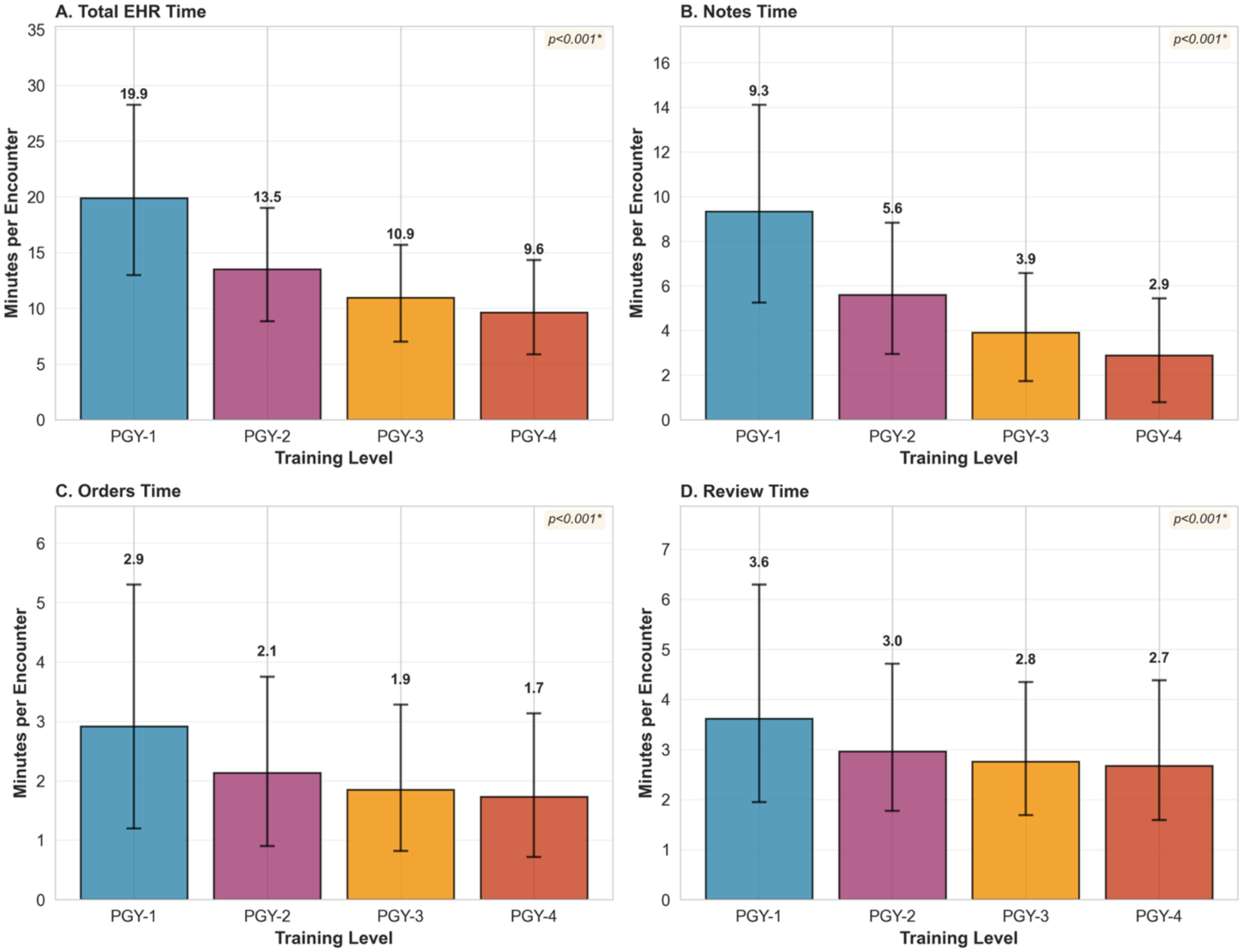
EHR Time per Encounter by Training Level. Median encounter-attributed EHR time (in minutes) by training level (PGY-1 to PGY-4) for (A) total EHR time, (B) Notes (documentation) time, (C) Orders (computerized provider order entry) time, and (D) Review (chart, laboratory, and imaging review) time. Bars represent medians; error bars represent interquartile range. Tracking board time is excluded from these panels and reported separately at the shift level (Table 2. Shift defined as 9 clinical hours; after-shift = activity occurring after the 9-hour clinical period. *P-values from Kruskal-Wallis tests on resident-level medians (one median per resident per PGY year) to account for within-resident clustering; see Methods.

**Table 2.**
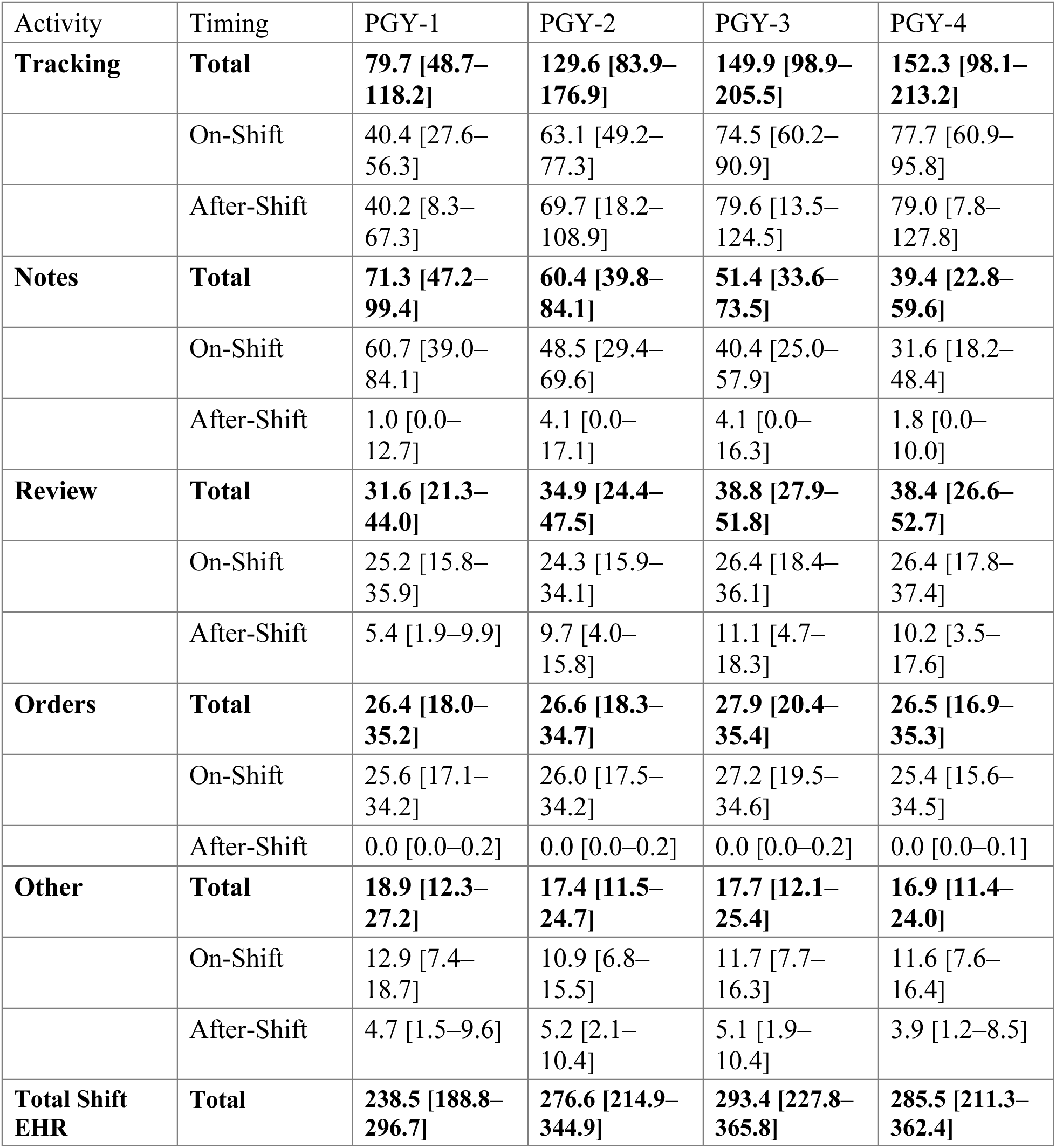
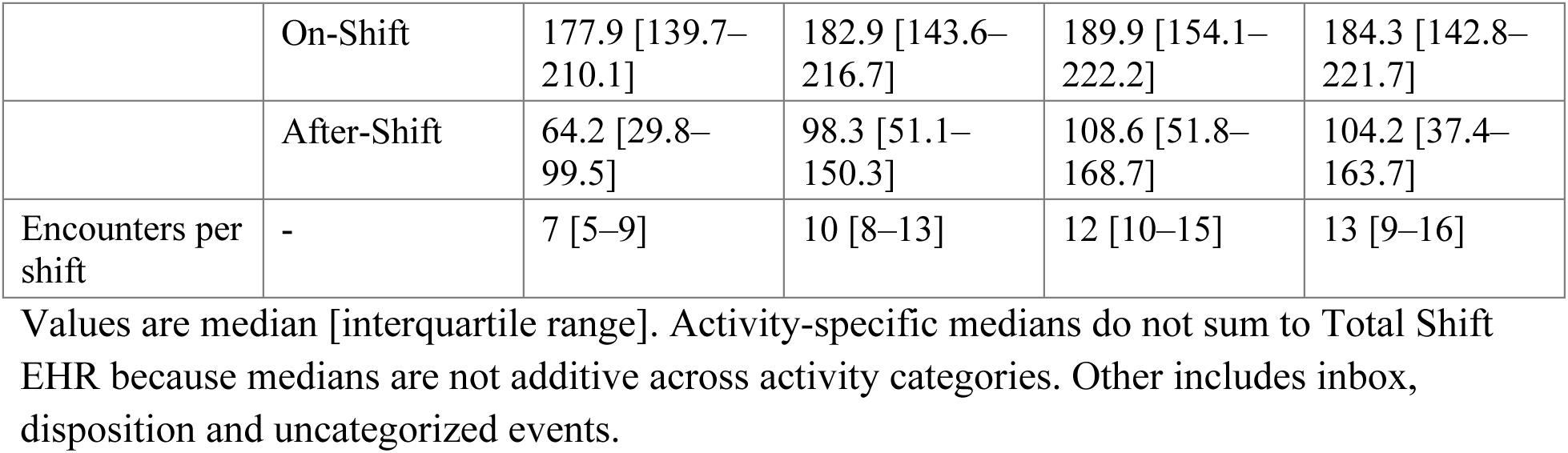
Per-Shift EHR Time by Activity and Timing.

| Activity | Timing | PGY-1 | PGY-2 | PGY-3 | PGY-4 |
| --- | --- | --- | --- | --- | --- |
| **Tracking** | **Total** | **79.7 [48.7–118.2]** | **129.6 [83.9–176.9]** | **149.9 [98.9–205.5]** | **152.3 [98.1–213.2]** |
|  | On-Shift | 40.4 [27.6–56.3] | 63.1 [49.2–77.3] | 74.5 [60.2–90.9] | 77.7 [60.9–95.8] |
|  | After-Shift | 40.2 [8.3–67.3] | 69.7 [18.2–108.9] | 79.6 [13.5–124.5] | 79.0 [7.8–127.8] |
| **Notes** | **Total** | **71.3 [47.2–99.4]** | **60.4 [39.8–84.1]** | **51.4 [33.6–73.5]** | **39.4 [22.8–59.6]** |
|  | On-Shift | 60.7 [39.0–84.1] | 48.5 [29.4–69.6] | 40.4 [25.0–57.9] | 31.6 [18.2–48.4] |
|  | After-Shift | 1.0 [0.0–12.7] | 4.1 [0.0–17.1] | 4.1 [0.0–16.3] | 1.8 [0.0–10.0] |
| **Review** | **Total** | **31.6 [21.3–44.0]** | **34.9 [24.4–47.5]** | **38.8 [27.9–51.8]** | **38.4 [26.6–52.7]** |
|  | On-Shift | 25.2 [15.8–35.9] | 24.3 [15.9–34.1] | 26.4 [18.4–36.1] | 26.4 [17.8–37.4] |
|  | After-Shift | 5.4 [1.9–9.9] | 9.7 [4.0–15.8] | 11.1 [4.7–18.3] | 10.2 [3.5–17.6] |
| **Orders** | **Total** | **26.4 [18.0–35.2]** | **26.6 [18.3–34.7]** | **27.9 [20.4–35.4]** | **26.5 [16.9–35.3]** |
|  | On-Shift | 25.6 [17.1–34.2] | 26.0 [17.5–34.2] | 27.2 [19.5–34.6] | 25.4 [15.6–34.5] |
|  | After-Shift | 0.0 [0.0–0.2] | 0.0 [0.0–0.2] | 0.0 [0.0–0.2] | 0.0 [0.0–0.1] |
| **Other** | **Total** | **18.9 [12.3–27.2]** | **17.4 [11.5–24.7]** | **17.7 [12.1–25.4]** | **16.9 [11.4–24.0]** |
|  | On-Shift | 12.9 [7.4–18.7] | 10.9 [6.8–15.5] | 11.7 [7.7–16.3] | 11.6 [7.6–16.4] |
|  | After-Shift | 4.7 [1.5–9.6] | 5.2 [2.1–10.4] | 5.1 [1.9–10.4] | 3.9 [1.2–8.5] |
| **Total Shift EHR** | **Total** | **238.5 [188.8–296.7]** | **276.6 [214.9–344.9]** | **293.4 [227.8–365.8]** | **285.5 [211.3–362.4]** |
|  | On-Shift | 177.9 [139.7–210.1] | 182.9 [143.6–216.7] | 189.9 [154.1–222.2] | 184.3 [142.8–221.7] |
|  | After-Shift | 64.2 [29.8–99.5] | 98.3 [51.1–150.3] | 108.6 [51.8–168.7] | 104.2 [37.4–163.7] |
| Encounters per shift | - | 7 [5–9] | 10 [8–13] | 12 [10–15] | 13 [9–16] |

Values are median [interquartile range]. Activity-specific medians do not sum to Total Shift EHR because medians are not additive across activity categories. Other includes inbox, disposition and uncategorized events.

### After-shift work patterns

At the encounter level, after-shift EHR work was present in 89.9–94.4% of encounters across all training levels (Figure 2B). The distribution of the percentage of an encounter’s EHR time occurring after-shift was right-skewed (Figure 2A), with median after-shift percentages of 9.8% in PGY-1, 16.1% in PGY-2, 17.5% in PGY-3, and 15.6% in PGY-4. Among encounters with any after-shift work, median duration was 1.9 minutes in PGY-1, 2.0 in PGY-2, 1.8 in PGY-3, and 1.4 in PGY-4 (Figure 2C). At the shift level, when after-shift EHR time was summed across all encounters in a shift, encounter-attributed after-shift time was a median of 16.2 minutes (IQR 6.8–33.7) for PGY-1 residents and 19.7 minutes (IQR 9.2–37.2) for PGY-4 residents (Table 2). After-shift tracking board time was an additional 40.2 minutes (IQR 8.3–67.3) for PGY-1 and 70.9 minutes (IQR 7.8–127.8) for PGY-4. Combined, total after-shift EHR time was a median of 64.2 minutes per shift for PGY-1 and 104.2 minutes for PGY-4 (p<0.001).

**Figure 2.**
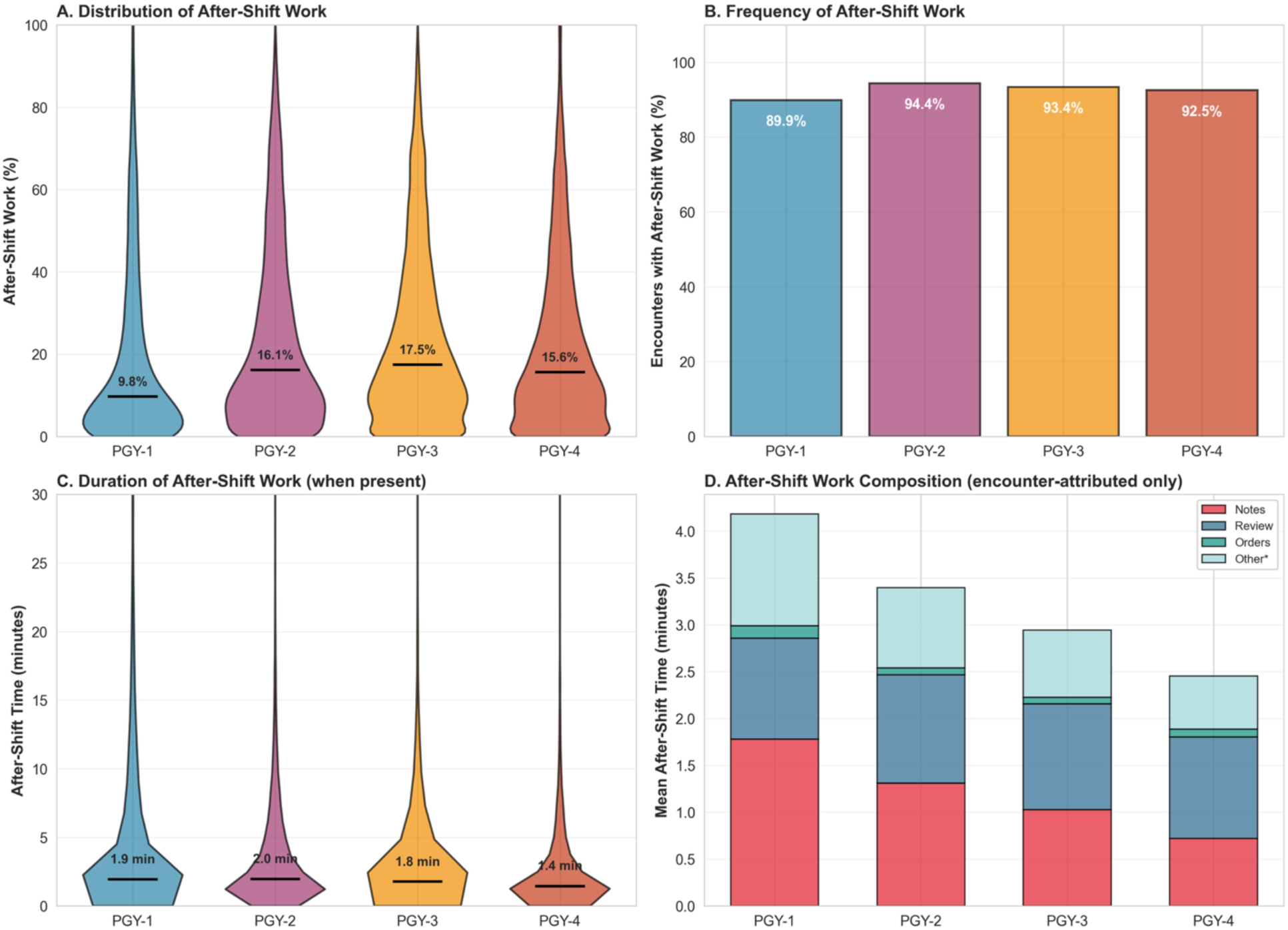
After-Shift Work Patterns by Training Level. After-shift work distribution (A), frequency (B), duration in minutes (C), and encounter-attributed composition (D), by training level (PGY-1 to PGY-4). Panel D shows mean values for encounter-attributed after-shift time only; tracking board time is excluded and reported separately at the shift level (Table 2). *Other includes inbox, disposition, best practice alerts, and miscellaneous EHR activities. Black line in violin plots = median. Shift defined as 9 clinical hours; after-shift = activity occurring after the 9-hour clinical period. P-values from Kruskal-Wallis tests on resident-level medians to account for within-resident clustering; see Methods.

Shift-level tracking board time (Table 2) dominated the after-shift burden across all PGY levels and increased with training, with total tracking per shift rising from a median of 79.7 minutes (IQR 48.7–118.2) in PGY-1 to 152.3 minutes (IQR 98.1–213.2) in PGY-4. On-shift tracking rose from 40.4 to 77.7 minutes per shift, and after-shift tracking rose from 40.2 to 79.0 minutes per shift, from PGY-1 to PGY-4. Within the encounter-attributed portion of after-shift work, notes and chart review accounted for the largest share, with the remaining time divided among orders, inbox management, disposition updates, and other activities (Figure 2D).

### Documentation efficiency and note composition

Documentation patterns varied across training levels. The majority of note content (approximately 81–84% of all characters) consisted of system-generated content and templates at every PGY level. The proportion of characters from manual entry decreased from 7.7% in PGY1 to 6.3% in PGY4. Dot phrase tool usage increased from 1.6% to 3.0%, and voice recognition increased from 6.3% to 9.0%.

Note volume and documentation speed changed over the course of training. Median note length decreased by 79% from 24,960 characters in PGY1 to 5,254 characters in PGY4 (p<0.001). Active input speed (typing and dictation combined) was relatively stable across training, ranging from 343 characters per minute in PGY-1 to 410 in PGY-3 and 356 in PGY-4.

### Training progression patterns

EHR time per encounter decreased progressively across training years, with the largest reductions occurring between PGY-1 and PGY-2 (median 19.9 to 13.5 minutes). Longitudinal analysis of 52 residents with complete data across all four PGY years confirmed within-person improvement: residents demonstrated a median 52.4% reduction in total EHR time per encounter from PGY1 to PGY4, and paired comparison using the Wilcoxon signed-rank test showed significant individual-level efficiency gains (p<0.001; Supplementary Figure 1).

### Pairwise comparisons across training levels

Group level differences across PGY remained statistically significant for total EHR time, notes time, orders time, and chart review time after aggregating to resident-level medians to account for within resident clustering (all p<0.001, Kruskal-Wallis; Supplementary Table 1). Pairwise comparisons revealed activity-specific learning trajectories. Note-writing time decreased at every adjacent PGY transition (all p<0.001). Order-entry efficiency improved most between PGY-1 and PGY-2 (p<0.001), with no significant adjacent-level differences seen thereafter. Chart review efficiency followed a non-monotonic pattern, with a significant reduction between PGY-1 and PGY-2 (p=0.003), no significant change between PGY-2 and PGY-3 (p=0.526), and a further significant reduction between PGY-3 and PGY-4 (p=0.017). At the shift level, encounter-attributed after-shift time did not differ significantly between adjacent senior PGY groups, while tracking-board time increased robustly between PGY-1 and PGY-3 (all pairwise p<0.001) before plateauing between PGY-3 and PGY-4 (p=0.262).

Scribe utilization ranged from 7.0% of encounters for PGY-1 residents to 27.2% for PGY-4 residents. Scribed encounters had modestly lower EHR time per encounter (median difference 0.7–2.3 minutes, p<0.001 for PGY 2–4), with the most pronounced reduction in note-writing time. However, the training-level gradient in EHR efficiency was preserved in both scribed and non-scribed encounters (all p<0.001, Kruskal-Wallis).

## DISCUSSION

In this multi-year audit-log study of EM residents, EHR work changed in two important ways across training: per-encounter time declined substantially, and documentation became more succinct, yet a large proportion of encounters still involved after-shift EHR activity. Residents thus became markedly more efficient on shift, particularly with documentation, while a nontrivial amount of work continued to occur after the scheduled end of shifts, most commonly in patient-tracking and flow-related tasks. Taken together, these patterns suggest that EHR proficiency develops throughout residency but that structured features of ED workflow are associated with computer-based work extending beyond scheduled clinical time.^15,17^

Our findings extend prior quantifications of trainee EHR burden. In EM, Olson and colleagues estimated that residents spend approximately 2,171 hours in the EHR over three years, with documentation and chart review comprising the largest shares.^14^ Objective studies in internal medicine and surgery similarly show that interns spend more time in the EHR and gradually improve efficiency over time.^18,19^ Longitudinal ambulatory data from residents in multiple specialties demonstrate that EHR time per patient decreases with experience while the fraction of time spent outside scheduled clinical hours often does not change.^15^ Survey data from a large national cohort of family medicine residents also show that high levels of “pajama time” are associated with lower knowledge scores, lower professional and training satisfaction, and higher burnout symptoms, underscoring that when EHR work occurs may matter as much as how long it takes.^20^ Our PGY-resolved, encounter-level analysis in a shift-based EM setting builds on this literature by showing that both patterns can coexist: per-encounter time decreases steeply, while after-shift work remains nearly universal and, in relative terms, may appear larger for more senior residents.

The per-shift view, however, qualifies the simple efficiency narrative. Although per-encounter EHR time decreased substantially across training, the *absolute* per-shift EHR burden actually increased from PGY-1 to PGY-3 (median 238.5 to 293.4 minutes) before plateauing in PGY-4. This pattern reflects a structural counterweight: per-encounter efficiency gains are partially offset by the increase in patient volume per shift. Activity-by-activity, notes are the only category in which individual level efficiency outpaces the volume increase, with per-shift documentation time falling from 71.3 to 39.4 minutes, despite seniors seeing nearly twice as many patients. For orders, review, and other categories, faster per-encounter performance is approximately balanced by greater encounter volume, leaving per-shift activity time stable or slightly increasing.

Tracking board time grew most prominently with seniority (79.7 to 152.3 minutes per shift), consistent with the expanding cross-patient coordination role of senior residents. Together, these patterns suggest that residency training produces real per-encounter efficiency gains, but those gains are absorbed by clinical productivity expectations, rather than translating into reduced total EHR exposure. The per-shift patient volumes observed here (median 7 encounters per shift for PGY-1 rising to 13 for PGY-4, corresponding to roughly 0.9 to 1.6 new patients per clinical hour) are broadly consistent with published benchmarks of EM resident productivity, supporting the generalizability of the workload context within which the observed efficiency gains occur.^21,22^

Importantly, the observed efficiency gains were not confounded by differential patient complexity across training levels. PGY-1 residents cared for patients with lower acuity and lower admission rates, suggesting that the reduced per-encounter EHR time among senior residents occurred despite—not because of—managing clinically comparable or slightly higher-acuity patients.

Our cluster-summary approach revealed that while the overall efficiency gains were robust to within-resident clustering, the learning trajectory varied by EHR activity type. Note writing efficiency differentiated at every PGY level. Order-entry efficiency improved most between PGY-1 and PGY-2, with no significant differences between PGY 2,3, and 4, suggesting rapid acquisition of ordering proficiency. Chart review efficiency followed a non-monotonic pattern with improvement between PGY-1 and PGY-2, a plateau between PGY-2 and PGY-3, and renewed improvement between PGY-3 and PGY-4. developed more gradually, with significant reduction emerging only at PGY-4. This pattern is consistent with a clinical learning trajectory in which documentation shortcuts and ordering proficiency develop before the more nuanced skill of efficient chart navigation and information extraction.

A key contribution of this study is characterizing how efficiency gains are distributed across EHR activities. Documentation time accounted for the largest share of improvement, and the pattern of change—shorter notes, faster input speed, stable template reliance—suggests that residents produce less text more quickly rather than simply accelerating their existing approach. Residents’ notes became shorter, manual and dictated text was produced more quickly, and system-generated content remained the dominant source of text. Virtually all residents demonstrated large within-person reduction in documentation time across training. These findings are consistent with prior work showing template optimization and “dotphrase” tools can shorten documentation time and that benefits plateau once basic adoption is achieved.^23^ They also align with work conceptualizing documentation as “composing,” in which the act of writing is tightly linked to clinical reasoning and learning for trainees.^24^ These findings are consistent with note design and tool fluency contributing to efficiency gains, though growing clinical expertise (pattern recognition, familiarity with clinical language, and comfort with diagnostic reasoning) likely also accelerates documentation. Our data cannot distinguish between these mechanisms, but together they suggest that documentation practice is a key domain of efficiency improvement during residency, while cautioning against approaches that remove trainees entirely from documentation and thereby risk undermining its educational value.

The composition and timing of after-shift work offer a complementary perspective on resident workload. Across PGY levels, more than 89% of encounters involved some after-shift activity, and the median proportion of EHR time occurring after-shift increased modestly over training. Most after-shift time was devoted to tracking-board and patient-flow activities rather than narrative documentation, with this share highest among senior residents. In the Epic EHR, tracking-board interactions include reviewing the ED patient list, checking result status, monitoring bed assignments and patient dispositions, and following up on pending orders—activities that reflect ongoing clinical responsibility for patients who remain in the department after a resident’s shift ends. Because our after-shift window begins after the additional “wrap up” hour, these activities likely represent residual monitoring of patients whose care was initiated during the shift rather than sign-out itself. Notably, these activities persist despite a dedicated additional hour after clinical care specifically allocated for sign out and documentation completion. This pattern is conceptually consistent with studies in inpatient and ambulatory settings showing lingering EHR activity for day teams beyond scheduled hours and stable proportions of “after-hours” work despite improved efficiency.^15,17^ It also reflects an evolving role shift from primary documenter to team and flow coordinator: junior residents complete more of their own charting after-shift, whereas senior residents increasingly carry out cross-patient coordination and tracking tasks that are less easily completed during high-intensity clinical periods. These activities are essential to department functioning yet are seldom captured in conventional descriptions of “documentation burden” and may be undercounted in duty hour monitoring.

These findings have several implications for graduate medical education. First, the documentation domain appears highly teachable and amenable to improvement with experience, suggesting it may be a productive target for early educational efforts. Early, explicit training in note parsimony, template personalization, and efficient use of dot phrase and dictation tools is likely to yield meaningful reductions in per-encounter documentation time, especially for interns.^23,25^ At the same time, documentation is an important vehicle for developing and demonstrating clinical reasoning and for generating artifacts used in assessment and feedback; qualitative work suggests that composing clinical notes plays a pivotal role in how trainees think and learn, particularly early in training.^24^ Efficiency initiatives should therefore focus on eliminating low-value redundancy rather than minimizing time or length, while preserving the trainee’s role in independently writing the medical decision-making section. Second, the growing share of tracking-board and flow work among senior residents suggests a need for equally intentional training in ED operations, team coordination, and handoff strategies, recognizing these activities as core competencies rather than invisible work around the margins of clinical care.

There are also system-level implications. Because a substantial fraction of after-shift time is devoted to tracking and flow rather than narrative documentation, interventions limited to documentation tools may have limited impact on after-shift burden. Team-based documentation support has been associated with reduced documentation time and EHR time, particularly after an adoption period, in large ambulatory cohorts.^1^ Similar models, or emerging ambient scribe tools, may reduce documentation burden for residents, but they are unlikely to address tracking-board work that is tightly coupled to real-time departmental operations.^26,27^ More broadly, as ambient scribe and AI-assisted documentation technologies become embedded in clinical training environments, future studies will need to examine how these tools reshape the EHR efficiency learning curve and what new documentation competencies they require of trainees.

Conversely, changes in staffing models, care coordination roles, or handoff processes that shift tracking responsibilities back into scheduled shifts could meaningfully reduce after-shift work even if per-encounter documentation time remained unchanged. From a monitoring standpoint, pairing audit-log metrics with measures of well-being and learning could help programs identify residents who accrue disproportionate after-shift work and evaluate whether interventions are improving both efficiency and trainee experience, in line with calls to connect EHR metrics, burnout, and training outcomes.^15,20^

## LIMITATIONS

This study has several limitations. It was conducted at a single, large academic medical center using a specific EHR configuration and residency structure, which may limit generalizability to other settings. Even within this setting, our analysis captures only EHR activity during emergency medicine shifts at the primary site; residents in our program complete substantial portions of their clinical training at other emergency departments and during off-service rotations, where their EHR activity is not included in our data. The resulting trajectories therefore reflect documentation efficiency at the home site rather than total EHR exposure, and skill acquisition occurring elsewhere may contribute to observed efficiency gains in ways we cannot directly measure. Additionally, we do not account for supervisory or sign out encounters, in which residents were heavily engaged in patient evaluation and clinical decision-making.

Vendor audit logs have known measurement limits: they provide objective data on user interactions but do not capture cognitive work, informal teaching, or brief offline activities, and misclassification of activity categories is possible despite careful mapping.^28^ In particular, brief documentation entries made through the ED tracking board interface cannot be separated from other tracking activity in the audit log, which may modestly inflate measured tracking time and underestimate documentation time, although this overlap is unlikely to alter the directional findings. The study period also spanned the COVID-19 pandemic, during which patient volumes, case mix, and operational pressures fluctuated and may have affected EHR use in ways that are difficult to disentangle from training effects. Finally, we did not directly measure resident well-being, clinical performance, or patient outcomes. Inferences about the consequences of EHR burden for learning or burnout therefore remain indirect and should be addressed in future work that links audit-log data to these outcomes, including work that has associated high after-hours EHR use with lower knowledge scores and higher burnout among residents.^15,20^

## CONCLUSION

In this multi-year audit-log study, EM residents became substantially more efficient in EHR use across training, particularly in documentation, yet after-shift EHR work remained common and was largely driven by tracking and flow-related activities. These findings suggest that both trainee-facing interventions and system-level changes will be needed to meaningfully reduce EHR burden during residency.

## Data Availability

All data produced in the present study are available upon reasonable request to the authors

## Notes

### Competing Interest Statement

The authors have declared no competing interest.

### Funding Statement

This study did not receive any funding

### Author Declarations

The Stanford University institutional review board approved the study (Protocol # 69107)

