## Supplementary Appendix for "Increasing Efficiency, Persistent Burden: Longitudinal Analysis of EHR Use and After-Hours Work in Emergency Medicine Residency"

Table of Contents

- Supplementary Figure 1: Individual Longitudinal EHR Efficiency Trajectories
- Supplementary Table 1: Clustered Statistical Testing Results
- Supplementary Table 2: Shift-Level Tracking Board Time
- Supplementary Methods: Additional Methodological Details

**Supplementary Figure 1: Individual Longitudinal EHR Efficiency Trajectories**

Individual-level progression of EHR efficiency across all four years of training for 52 residents with complete longitudinal data. Panel A displays resident-specific trajectories (gray lines) of median EHR time per encounter by PGY level, with population median (red circles/line) and mean (blue squares/line) overlaid. Both summary trajectories demonstrate a consistent downward trend in EHR time per encounter from PGY1 to PGY4. Panel B shows the distribution of within-person percentage improvement from PGY1 to PGY4. The vast majority of residents (46/52, 88.5%) demonstrated reduced EHR time per encounter, with a median improvement of 52.4%. A paired comparison of PGY1 vs PGY4 median per-encounter EHR time using the Wilcoxon signed-rank test confirmed significant within-person efficiency gains (p < 0.001).

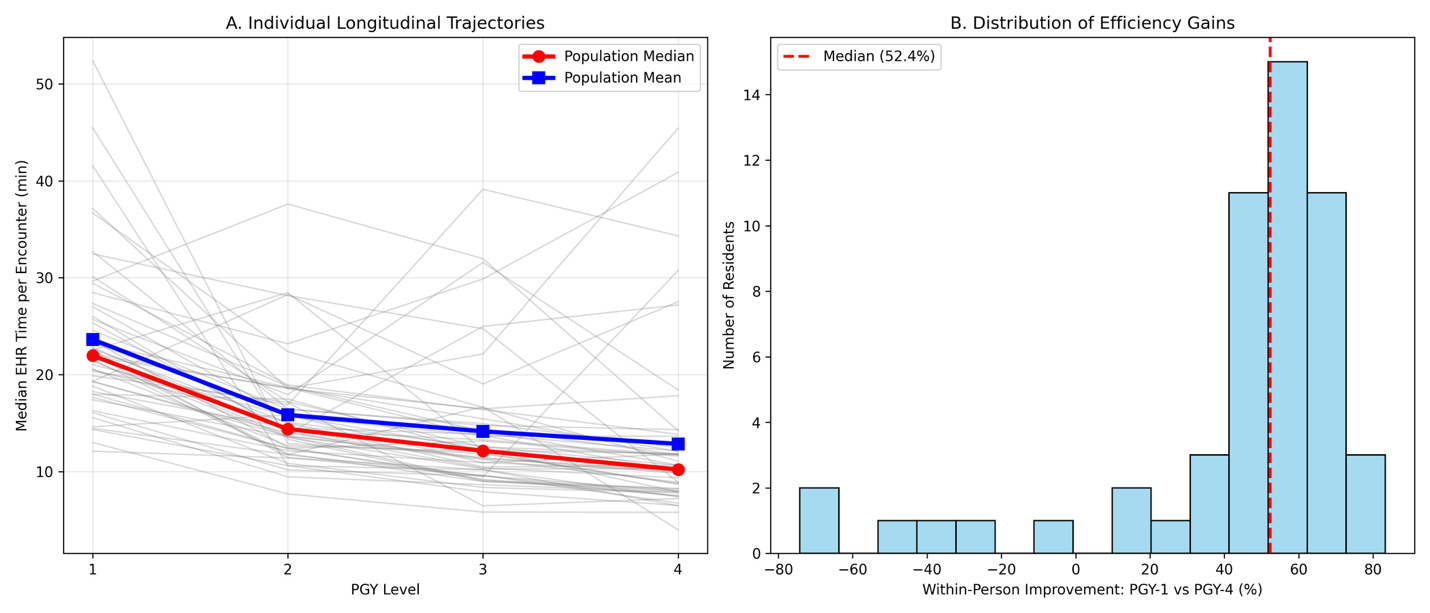

**Supplementary Table 1: Clustered Statistical Testing Results**

Because encounters are nested within residents, EHR-time measures were aggregated to the resident level (one median per resident per PGY year) before formal hypothesis testing. This cluster-summary approach treats each resident as a single observation within each training year, eliminating the inflated precision that would result from treating 167,010 encounters as independent observations.

All primary EHR time metrics remained statistically significant (p<0.001) after resident-level aggregation.

**Statistical methods**

Kruskal-Wallis tests on resident-level medians were used for omnibus comparisons. Mann-Whitney U tests (two-sided) were performed for pairwise comparisons. This approach makes no distributional assumptions and eliminates inflated precision from non-independent encounters. P-values are presented unadjusted; interpretation emphasizes magnitude and direction of effects alongside statistical significance.

| Metric | PGY-1 | PGY-2 | PGY-3 | PGY-4 | KW p-value | 1 vs 2 | 1 vs 3 | 1 vs 4 | 2 vs 3 | 2 vs 4 | 3 vs 4 |
| --- | --- | --- | --- | --- | --- | --- | --- | --- | --- | --- | --- |
| **Total EHR Time (min)** | 22.1 (18.6-28.5) | 15.3 (12.4-18.6) | 12.5 (10.1-15.5) | 10.2 (8.2-12.1) | <0.001 | <0.001 | <0.001 | <0.001 | <0.001 | <0.001 | <0.001 |
| **Notes Time (min)** | 10.3 (7.5-13.1) | 5.4 (4.0-7.2) | 4.4 (3.1-5.5) | 3.0 (2.3-4.2) | <0.001 | <0.001 | <0.001 | <0.001 | <0.001 | <0.001 | <0.001 |
| **Orders Time (min)** | 3.0 (2.3-3.7) | 2.0 (1.7-2.4) | 1.8 (1.5-2.1) | 1.8 (1.5-2.4) | <0.001 | <0.001 | <0.001 | <0.001 | 0.001 | 0.142 | 0.178 |
| **Review Time (min)** | 4.3 (3.3-7.0) | 3.2 (2.5-5.5) | 3.1 (2.3-4.5) | 2.7 (2.1-3.4) | <0.001 | 0.003 | 0.001 | <0.001 | 0.526 | 0.001 | 0.017 |
| **Inbox Time (min)** | 0.5 (0.3-1.0) | 0.3 (0.1-0.8) | 0.2 (0.1-0.5) | 0.1 (0.0-0.3) | <0.001 | 0.032 | <0.001 | <0.001 | 0.080 | <0.001 | 0.002 |
| **After-Shift Time (min)** | 2.4 (1.2-5.1) | 2.2 (1.3-4.4) | 2.1 (1.2-4.9) | 1.3 (0.9-2.5) | <0.001 | 0.918 | 0.886 | 0.001 | 0.985 | <0.001 | <0.001 |
| **After-Shift (%)** | 12.8 (5.9-23.3) | 19.6 (11.7-30.3) | 22.4 (13.1-35.7) | 15.9 (11.3-26.1) | <0.001 | 0.001 | <0.001 | 0.012 | 0.084 | 0.279 | 0.005 |
| **Total Characters** | 25k (19.7k-29.1k) | 18.6k (15.1k-22.6k) | 15.6k (12.3k-18.2k) | 5.5k (2.3k-11.4k) | <0.001 | <0.001 | <0.001 | <0.001 | <0.001 | <0.001 | <0.001 |
| **Manual Characters** | 1664 (604-3055) | 451 (116-1280) | 223 (56-716) | 116 (10-503) | <0.001 | <0.001 | <0.001 | <0.001 | 0.005 | <0.001 | 0.017 |
| **Shift Enc-After** | 15.8 (9.3-25.6) | 19.1 (13.8-29.9) | 21.8 (15.5-38.4) | 18.7 (13.5-32.5) | 0.003 | 0.016 | <0.001 | 0.037 | 0.149 | 0.769 | 0.104 |
| **Shift Tracking** | 43.1 (26.0-54.7) | 68.7 (56.8-82.3) | 80.7 (68.2-97.7) | 78.1 (64.5-93.8) | <0.001 | <0.001 | <0.001 | <0.001 | <0.001 | 0.007 | 0.262 |
| **Shift Combined** | 61.7 (46.6-79.6) | 91.8 (75.9-118.7) | 113.7 (88.6-133.3) | 103.8 (84.0-125.2) | <0.001 | <0.001 | <0.001 | <0.001 | <0.001 | 0.023 | 0.123 |

**Supplementary Table 2: Shift-Level Tracking Board Time**

Tracking board time — time spent in the ED patient tracking board, census screen, and patient flow management displays — was analyzed as a shift-level metric because 97% of tracking board activity in the audit logs had no encounter-level attribution. Total tracking board time per shift is reported, stratified by on-shift and after-shift periods. Values are median (IQR) in minutes per shift.

Total tracking time per shift increased with training level (median 79.7 min/shift for PGY-1 to 152.3 min/shift for PGY-4), with after-shift tracking time rising from 40.2 to 79.0 min/shift across PGY-1 through PGY-4.

| Metric | PGY-1 (1,899) | PGY-2 (4,494) | PGY-3 (5,732) | PGY-4 (3,261) |
| --- | --- | --- | --- | --- |
| Total tracking/shift (min) | 79.7 (48.7–118.2) | 129.6 (83.9–176.9) | 149.9 (98.9–205.5) | 152.3 (98.1–213.2) |
| On-shift tracking (min) | 40.4 (27.6–56.3) | 63.1 (49.2–77.3) | 74.5 (60.2–90.9) | 77.7 (60.9–95.8) |
| After-shift tracking (min) | 40.2 (8.3–67.3) | 69.7 (18.2–108.9) | 79.6 (13.5–124.5) | 79.0 (7.8–127.8) |

Combined after-shift p<0.001 (clustered KW).

**Supplementary Methods: Additional Methodological Details**

Activity categorization schema

EHR activities were classified into seven categories using Epic audit-log metadata and activity descriptors, aligned with the main Methods section:

1. *Notes* – Documentation activities including history and physical notes, progress notes, discharge summaries, addenda and amendments, and note review and co-signature activities.
2. *Tracking* – Patient-flow management, including ED tracking board interactions, patient status updates, location changes, and disposition planning workflows.
3. *Review* – Chart review activities, including laboratory results review, radiology and imaging review, prior visit documentation review, medication reconciliation review, and consultant note review.
4. *Orders* – Computerized provider order entry, including medication orders, laboratory test orders, imaging orders, procedure orders, and consult requests.
5. *Disposition* – Care transition activities, including admission orders, discharge instructions, transfer workflows, and referral documentation.
6. *Inbox* – Communication and messaging activities, including in-basket message review and response, secure messaging, results notifications, and task management.
7. *Other* – Miscellaneous EHR activities not classified above, including null or uncategorized events and interactions with clinical decision support tools (for example, Best Practice Alerts).

Temporal window definitions

On-shift time was defined as EHR activity occurring from the scheduled shift start through 9 hours (the end of the clinical period). After-shift time was defined as EHR activity occurring after the 9-hour clinical period through 48 hours post-shift start. The 48-hour capture window ensured that delayed documentation was attributed to the originating shift.

Tracking board time — time spent viewing or interacting with the ED patient tracking board, census screen, and patient flow management displays — was analyzed as a shift-level metric. In the Epic audit log data, 97% of tracking board activity had a NULL patient encounter identifier, indicating it was not associated with a specific patient's chart. Tracking board time was therefore reported as total minutes per shift, stratified by on-shift and after-shift periods.

Activity attribution assigned each second of encounter-attributed EHR activity to the shift during which the associated encounter was active. Activity within the 9-hour clinical period was classified as 'on-shift,' and activity after the 9-hour mark was classified as 'after-shift.'

Exclusion criteria rationale

*Encounter Inclusion via Credit Table.* Encounter inclusion was based on treatment team assignment, using the institutional encounter credit logic. Each encounter was attributed to the resident formally signed up to the patient in the treatment team prior to initial disposition. This ensures that EHR activity is attributed only to the primary physician responsible for the bulk of documentation.

*Supervisory encounter exclusion.* Senior residents (PGY3–4) often work in supervisory roles when junior residents (PGY1–2) are concurrently assigned to the same patient. When both a junior and senior resident were credited on the same encounter, the senior resident’s record was excluded as the senior’s role was primarily supervisory. Excluding supervisory encounters for seniors prevents underestimation of senior-level EHR time and enhances comparability across PGY levels. Junior-resident encounters were retained in full. Particular PGY-4 shifts that include multi-team oversight were also excluded as these represent supervisory roles with minimal direct patient care.

Data quality and validation

Epic audit logs and active-use logs capture user interactions with timestamps at sub-minute resolution. Zero values for specific activity categories were treated as true zeros rather than missing data. Encounters with zero total EHR time were rare (<0.1%) and were excluded. Note-character data were available for 98.7% of encounters in the primary analysis; encounters missing note metrics were excluded only from documentation-specific summaries.

No arbitrary upper cutoffs were applied to time measures. Extreme values above the 99th percentile were reviewed and retained if clinically plausible (for example, very long ED stays). The skewed nature of the data motivated the emphasis on medians in the primary manuscript, with means reported in supplementary tables for completeness.

Statistical power considerations

The study included 167,010 encounters from 144 residents across 15,386 shifts. At the resident level (the unit of analysis for hypothesis testing), each PGY group contained sufficient resident-level observations to provide adequate power to detect clinically meaningful differences.
